# A Composite Ranking of Risk Factors for COVID-19 Time-To-Event Data from a Turkish Cohort

**DOI:** 10.1101/2022.01.05.22268765

**Authors:** Ayse Ulgen, Sirin Cetin, Meryem Cetin, Hakan Sivgin, Wentian Li

## Abstract

Having a complete and reliable list of risk factors from routine laboratory blood test for COVID-19 disease severity and mortality is important for patient care and hospital management. It is common to use meta-analysis to combine analysis results from different studies to make it more reproducible. In this paper, we propose to run multiple analyses on the same set of data to produce a more robust list of risk factors. With our time-to-event survival data, the standard survival analysis were extended in three directions. The first is to extend from tests and corresponding p-values to machine learning and their prediction performance. The second is to extend from single-variable to multiple-variable analysis. The third is to expand from analyzing time-to-decease data with death as the event of interest to analyzing time-to-hospital-release data to treat early recovery as a meaningful event as well. Our extension of the type of analyses leads to ten ranking lists. We conclude that 20 out of 30 factors are deemed to be reliably associated to faster-death or faster-recovery. Considering correlation among factors and evidenced by stepwise variable selection in random survival forest, 10∼15 factors seem to be able to achieve the optimal prognosis performance. Our final list of risk factors contain calcium, white blood cell and neutrophils count, urea and creatine, d-dimer, red cell distribution widths, age, ferritin, glucose, lactate dehydrogenase, lymphocyte, basophils, anemia related factors (hemoglobin, hematocrit, mean corpuscular hemoglobin concentration), sodium, potassium, eosinophils, and aspartate aminotransferase.

## Introduction

The purpose of meta analysis is to combine information from different datasets in multiple studies in order to provide robust and consistent conclusions on the effect of a factor on an outcome (Borenstein et al., 2009; Haidich, 2010). However, it is less common to attempt multiple analyses on the same set of data to extract robust information. For example, in investigating risk factors for COVID-19 infection susceptibility, disease severity (e.g. hospitalization), and mortality (Guan et al., 2020), the most common approach is to carry out one test to obtain p-value (J Tian et al., 2020; X Liu et al., 2020; Rosenthal et al., 2020; Williamson et al., 2020; Fadl et al., 2021; T Tian et al., 2020). The test can be t-test/Wilcoxon test for continuous variable, or *χ*^2^-test/Fisher’s test for discrete variables, with the value of a risk factor in samples within two groups compared. Alternatively, uni-variable logistic regression can be used, and the null hypothesis of regression coefficient to be zero is tested. For time-to-event data (survival data), the time from hospitalization to death of a COVID-19 patient can be used to examine which factor contributes to a faster death (per unit time rate of death), which can be done by the Cox regression (proportional hazard model). The null hypothesis of zero regression coefficient is then tested.

In this paper, we extended the above common practice in three directions, on a COVID-19 patient time-to-event data. The first is to use both p-value based measures and prediction performance based ones. Although p-value-based approach has advantages: the meaning is easy to understand and the result is easy to report, it also has problems. P-value itself, often treated as the “gold standard for statistical validity”, may not be so golden (Nuzzo, 2014). A change in true prior probability of a signal will change the prediction error even when the p-value is the same (Nuzzo, 2014; Colquhoun, 2017). There have already been proposals of alternatives for p-value in evaluating variables (Lu and Ishwaran, 2018; Halsey, 2019).

The second extension is to use multiple-variable methods as well as single-variable ones. Single-variable methods evaluate a variable in isolation with respect to other variables. As a result, they would not detect conditional importance of a variable and its interaction with other variables. Inconsistency, or larger confidence interval, between different datasets concerning the importance of a factor may well reflect the contextual heterogeneity in other variables (Ghahramani et al., 2020; Kermali et al., 2020). The multi-variable statistical/machine learning models (Strobl et al., 2008) are ideal to supplement the single-variable analyses, but are less mainstream in the case of COVID-19 data analysis, are most focused on prediction and diagnosis (An et al., 2020; Li et al., 2020; McCoy et al., 2021; Li et al., 2021; Bennett et al., 2021; Karthikeyan et al., 2021; Aljameel et al., 2021; Mahdavi et al., 2021; Cornelius et al., 2021; Kocadagli et al., 2022; Malik et al., 2022), not on variable evaluation such as in our work. There are more applications of machine learning and artificial intelligence in the context of COVID-19, ranging from drug repurposing to medical assistance (Zeng et al., 2020; Deepthi et al., 2021; Chen et al., 2022; Alafif et al., 2021; Khan et al., 2021; Piccaialli et al., 2021; Dogan et al., 2021; Majeed and Hwang, 2022). On the other hand, over-interpreting specific multi-variable models (Yan et al., 2020) might not be a good practice as it may not be applicable to other data (Barish et al., 2021).

The third extension is specific for time-to-event data. For our inpatient data, not only have we deceased patients admission-to-death time information, but also we have larger number of patients who are completely recovered and released. In a typical survival analysis, these patients’ time-to-release information would be treated as right-censored data. However, treating them as the main event of interest, extra information might be obtained (Cetin et al., 2021b,c).

With our three extensions, we are able to carry out ten analyses on the same set of data. Combining these analyses to get a composite ranking of risk factors for COVID-19 faster death or faster recovery, we effectively run a meta-analysis on the same dataset. Besides the standard testing for unit hazard ratio from Cox regression (thus p-value based), we also used single-variable random survival forest (Breiman, 2001; Ishwaran et al., 2008) (thus prediction performance based), multi-variable random survival forest and variable selection in regularized regression (thus multiple variable based). Two different measures of performance (discordance index and integrated Brier score) in single-variable random survival forest are used. Then all these analyses were repeated for the time-to-release data, resulting in ten different sets of results. We will show that our composite ranking of ten runs result in a more robust list of risk factors for COVID-19 severity than the p-value based method alone, and our selected factors are fully validated by being consistent with the literature.

## Methods and Data

### COVID-19 patient data

Our COVID-19 patient data was collected from Tokat State Hospital (Turkey), with 3084 people and 35 potential risk factors. This study was carried out with the approval of Gaziosmanpaşa University Faculty of Medicine Non-Interventional Clinical Research Ethics Committee (decision No: 22-KAEK-051). The 2682 outpatients do not have time-to-event data and would not be used in our survival analysis by RSF. For the remaining 402 inpatients, five factors have too much missing data (activated partial thromboplastin clotting time (aPTT), red blood cell (RBC) count, HbA1C, fibrinogen, and C-reactive protein (CRP)) and are not used. The remaining 30 factors are: age, gender, glucose, d-dimer, calcium, chloride, potassium, sodium, creatine, ferritin, urea, alanine aminotransferase (ALT), aspartate aminotransferase (AST), lactate dehydrogenase (LDH), white blood cell (WBC) or leukocyte count, neutrophils (NEU) cell count, lymphocyte (LYM) cell count, monocyte (MON) cell count, eosinophils (EOS) cell count, basophils (BAS) cell count, platelets (PLT) cell count, hemoglobin (HGB) count, hematocrit (HCT), mean corpuscular volume (MCV), mean corpuscular hemoglobin (MCH), mean corpuscular hemoglobin concentration (MCHC), red cell distribution widths (RDW.CV and RDW.SD), mean platelet volume (MPV), platelet distribution width (PDW). Some of these factors still have missing data, but none of the missing rate exceeds 14%.

### Assessing independent variables in their contribution to time-to-event dependent variable using four methods

Our data is of the following form: 30 independent variables, 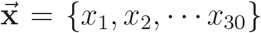, and one dependent variable 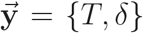, where T is the time to the event, and event status *δ* can take the value of 1 (event 1, or death), 2 (event 2, or release from hospital), 0 (right censored). We do not have any samples with right-censored time. How to handle multiple events data is usually within the scope of “competing risks” survival analysis (example: death from heart attack versus death from stroke); though in our case, the two events, risk and benefit, do not point to the same direction. How to analyze our special type of data will be discussed in the Result section. Here we simply reset *δ* = 2 to *δ* = 0 and summarize the existing methods.

All four of our analyses aim at finding which independent variable is associated with the dependent variable. We will describe each method as below. (1) In Cox regression, instead of modelling and fitting the survival function, an arbitrary baseline survival function is assumed, and a change in independent variable is assumed to lead to a constant multiple of the whole baseline curve (proportional hazard hypothesis):

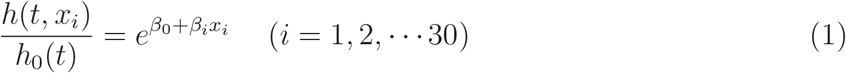

where *h*_0_(*t*) is the arbitrary baseline hazard function, *h*(*t, x*_*i*_) is the hazard function with one of the independent variable present. The p-value *p*_*i*_ for testing *β*_*i*_ = 0 measures the statistical significance of the contribution of *x*_*i*_ to the time-to-event data, and 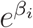 measures the hazard ratio with one unit change in *x*_*i*_.

(2) Random survival forest (RSF) (Ishwaran et al., 2008) is an extension of Random Forest (RF) (Breiman, 2001) to handle time-to-event data. RF/RSF construct many decision trees (therefore “forest”) that separate the dependent variable value in two daughter nodes as much as possible (for introduction on RF, see, for example, (Louppe, 2014; Fernández-Delgado, et al. 2014)). Once the splitting of nodes in a tree is done, in each external node, the cumulative hazard function (CHF) can be estimated by the Nelson-Aalen estimator (Ishwaran et al., 2008). The CHF with an independent variable *x*_*i*_ can be obtained by tracing the path on the tree, according to the *x*_*i*_ value, to reach the external node (Ishwaran et al., 2008):

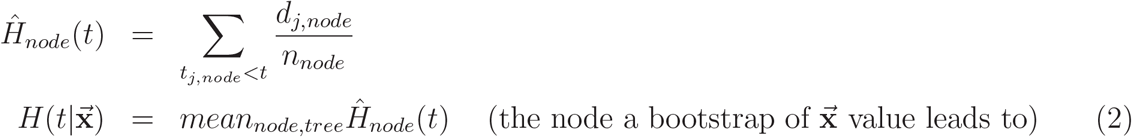

The performance of a RSF is measured by samples not used in the construction of the tree/forest, as only 1−*e*^−1^ ≈ 63.2% of the data are used in a sample-with-replacement approach (bootstrap), and those are called out-of-bag (OOB) samples. The CHF of individual OOB samples can be obtained in a similar way by tracing the path along a tree by its independent variable values, and averaged over trees (Ishwaran et al., 2008). Averaging over all OOB samples lead to an ensemble prediction of the CHF. As all samples will become an OOB in one of run of a tree, if the number of runs is large enough, a RSF will produce a predicted CHF for each sample.

Using the predicted CHF from the RSF program, two errors can be used to measure the prediction performance. One is Harrell’s concordance index (C-index) (Harrell Jr., 1982) whose complement can be called discordance index (D-index) (Cetin et al., 2021a). The C-index (D-index) is simply the number of sample pairs whose event time and predicted CHF are consistent (not consistent), divided by the number of permissible sample pairs:

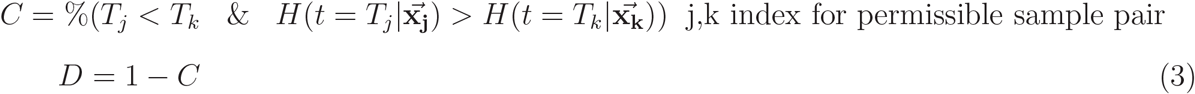

The second measure is (integrated) Brier score (IBS) (Brier, 1950), which is the squared difference between the actual value (e.g. binary indicator for survival) and the predicted value (e.g., predicted survival probability *Prob*(*T* > *t*)), integrated over available time-to-event points in the sample:

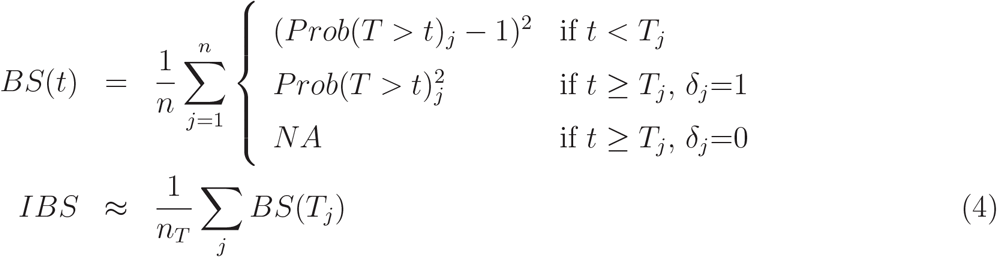

where *n*_*T*_ is the number of samples with *δ* = 1 and the sum is over these samples.

(3) RF/RSF also provides a performance-based evaluation of individual variables when all variables 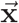 are used in RF/RSF (Breiman, 2001; Ishwaran et al., 2008). We refer to the permutation based measure of variable important as VIM (the other choices are external node purity based, such as the Gini index (Nembrini et al., 2018)). In this approach, a variable is removed (version-1) or randomized by permuting its value among samples (version-2), and the RF/RSF performance before and after permutation is compared:

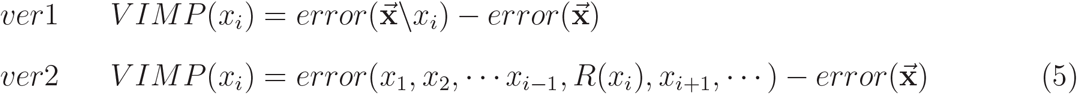

where R() refers to the random permutation step. Usually, it is the version-2 that is implemented in the commonly used programs. The variable that leads to the largest decrease of performance (or largest increase of error) is the most important variable. Note that this approach provides only a ranking of variables, and no cutoff of the list to separate important from unimportant variables is given.

(4) The regularized (Hastie et al., 2009) regression LASSO (Tibshirani, 199b, 1997) for Cox regression for multiple variables is:

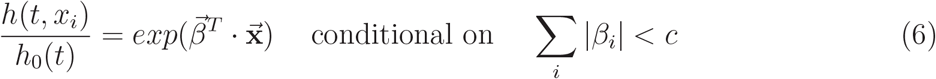

When *c* → 0, all regression coefficients are zero. As *c* increases, {*β*_*i*_} gradually emerge from zero to non-zero value one by one. It is a variable selection technique, and the order of variables being selected can be used to rank them. For example, the first variable with non-zero coefficient is ranked no.1. A LASSO plot (how regression coefficient values change with the constraint) can be used to easily find the rank order.

### Programs used

The R statistical platform (https://www.r-project.org) is used for our analysis. We use the R package *randomForestSRC* for random survival forest (Ishwaran and Kogalur, 2007), our own R functions for calculating IBS errors (http://github.com/wlicol/coxrsf/) (Cetin et al., 2021a), *survival* package for Cox regression and other survival analyses (Therneau and Grambsch, 2010). For LASSO on right-censored data, we use the R package *glmnet* (Simon et al., 2010). The Kendall correlation and testing of zero correlation is carried out by the R function *cor(* … *method=“kendall”)* and *cor*.*test(* … *method=“kendall”)*.

In the main RSF program we used, the *rfsrc* function from *randomForestSRC* package, the default number of trees (*ntree*=1000), default number of variables for splitting a branch 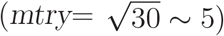, default minimum (average) number of samples in the external nodes (*nodesize*=15), are used. We choose samptype = “swr” (sampling with replacement), na.action = “na.impute” (imputing the missing value). The decision to use several default parameter settings is based on our experimenting with the parameter values (Probst et al., 2019), as have been done previously (Cetin et al., 2021a). Note that sampling with replacement is not the default setting in *rfsrc*, but consistent with the original proposal by Breiman.

## Results

### Survival analyses with two types of mutually exclusive events

Our time-to-event data contains two events of very different nature. As early as 1980s, it was suggested to use a type-specific (cause-specific) use of standard survival analysis program by switching the second type event to right-censored event (i.e., convert *δ*=2 to *δ*=0) (Kalbfleisch and Prentice, 1980). The cause-specific (cs) hazard is defined as

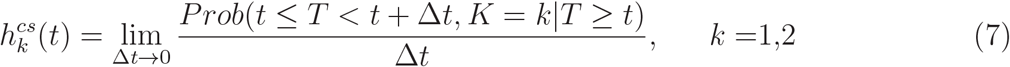

Although this definition clearly points to the possible cause-specific hazard for the second event, most people only concern about the main event, death, and do not consider the second event type.

We argue here that to run a survival analysis on the time-to-release data, while masking death as right-censored, actually provide valuable information. If 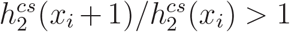, then a higher value of *x*_*i*_ leads to a faster release of a patient, and the variable-*i* is protective. For that same variable, we would expect 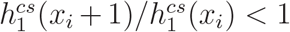 for the death event. The deceased samples should be more important in estimating 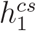. Similarly, the released samples should be more useful for the estimation of 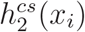. Therefore, we believe running the same survival analysis twice to find variables contributing to the two cause-specific hazard-ratios would use the dataset more fully.

In competing risk survival analysis, there is another population approach called Fine-Gray model (Fine and Gray, 1999) which defines the following subdistribution hazard:

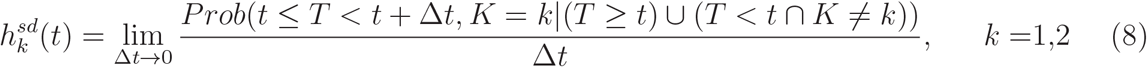

where the occurring of a type-2 event has no impact on the calculation of 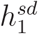, with the underlying assumption that the sample experiencing type-2 event may continue to experience a type-1 event. However, this scenario is impossible in our example because our two types of events, release from hospital and die from COVID-19, are mutually exclusive. Using cause-specific survival analysis for mutually exclusive events is explicitly recommended in (Allison, 2014).

### Composite ranking of factors based on five measures (time-to-death)

The factors are ranked five times using five measures: D-index from single-variable RSF, IBS from single-variable RSF, p-value for testing unit hazard ratio by single-variable Cox regression, permutation based VIM from the full model RSF, the variable selection order in LASSO. We found that for full-model RSF, because of random components in a run (both samples and variables are randomly chosen in a tree formation), the ranking order may change from run to run, especially for low-ranking factors. Therefore, we run the full-model RSF 100 times and the average of these runs is used.

Table 1 shows the results of these five measures. The rank for each method is within the parentheses, and the factors are listed by the overall rank order. It can be seen that some factors are consistently ranked high no matter what method is used (e.g., urea is ranked no.1 by all five methods, neutrophils and calcium are ranked within top 5 by all five methods). Other factors are not ranked consistently among methods: e.g., LDH, AST are ranked high in the three RSF based methods but ranked lower by Cox and LASSO; sodium is ranked lower in single-variable RSF measures; etc. The consistency is reassuring that the very top factors would be discovered by any method. The inconsistency or variation provides a basis for our approach of combining multiple rankings to improve the robustness of the result, even for one dataset.

**Table 1:** Composite ranking of factors based on time-to-death of COVID-19. Column 1 (Col-1): composite rank from five analyses; Col-2: names of factor; Col-3: two error measures of single-variable random survival forest (RSF): discordance index (D) and integrated Brier score (IBS). The number in the parenthesis is the rank (also for Cols 4-6); Col-4: p-value from single-variable Cox regression; Col-5: permutation-based variable importance (VIM) from the full RSF model, averaged over 100 runs; COl-6: (ranking according to the variable selection order from LASSO; T for tied rank).

| combined rank<br>(time-to-death) | var | single-var RSF<br>D(rank), IBS(rank) | Cox<br>pv(rank) | C full RSF<br>VIM$\times 10^3$ (rank) | LASSO<br>(rank) |
| --- | --- | --- | --- | --- | --- |
| 1 | urea | .186 (1), .121 (1) | 1.4E-19 (1) | 38.2 (1) | (1) |
| 2 | calcium | .220 (3), .131 (3) | 5.6E-14 (4) | 12.6 (2) | (3) |
| 3 | NEU | .235 (4), .141 (5) | 3.1E-16 (2) | 9.05 (4) | (2) |
| 4 | WBC * | .252 (5), .153 (8) | 2.2E-14 (3) | 5.05 (8) | (7) |
| 5 | creatinine * | .214 (2), .127 (2) | 2.1E-12 (5) | 10.6 (3) | (19) |
| 6 | LYM | .326 (12), .167 (14) | 1.3E-9 (8) | 5.3 (7) | (5.5T) |
| 7 | d-dimer | .3 (8), .167 (16) | 1.1E-9 (7) | 1.13 (15) | (4) |
| 8 | glucose | .281 (7), .154 (9) | 4.4E-5 (14) | 2.69 (10) | (11) |
| 9 | sodium | .367 (18), .18 (21) | 4.2E-11 (6) | 2.6 (9) | (5.5T) |
| 10 | BAS | .377 (20), .14 (4) | 2.4E-6 (13) | 0.98 (17) | (9T) |
| 11 | chloride | .35 (14), .175 (20) | 4.24E-8 (9) | 1.32 (12) | (9T) |
| 12 | RDW.SD | .313 (10), .171 (18) | 5.8E-8 (10) | .56 (19) | (9T) |
| 13 | LDH | .26 (6), .15 (7) | .015 (23) | 7.56 (5) | (26) |
| 14 | AST | .302 (9), .15 (6) | .021 (24) | 5.54 (6) | (24) |
| 15 | EOS | .365 (17), .164 (11) | 4E-4 (17) | 1 (16) | (13.5T) |
| 16 | potassium | .319 (11), .167 (15) | 5.7E-4 (18) | 0.279 (23) | (15) |
| 17 | age | .373 (19), .171 (17) | 8.7E-8 (11) | 0.319 (21) | (13.5T) |
| 18 | ALT * | .354 (15), .165 (13) | .0403 (27) | 0.454 (20) | (12) |
| 19 | RDW.CV * | .335 (13), .165 (12) | 1.7E-4 (16) | 0.29 (22) | (25) |
| 20 | ferritin | .38 (22), .183 (22) | 3.6E-7 (12) | 0.186 (24) | (21) |
| 21 | MPV | .378 (21), .192 (24) | 1.5E-4 (15) | 0.0648 (25) | (16) |
| 22 | PDW | .356 (16), .174 (19) | .036 (25) | -0.0178 (26) | (21) |
| 23 | HCT | .474 (27), .214 (29) | .008 (22) | 1.16 (14) | (17) |
| 24 | MCHC | .407 (23), .193 (25) | .0011 (19) | -0.047 (27) | (18) |
| 25 | PLT | .443 (25), .2 (26) | .0019 (21) | 0.88 (18) | (23) |
| 26 | HGB * | .416 (24), .206 (27) | .0014 (20) | 1.22 (13) | (29) |
| 27 | MON | .475 (28), .208 (28) | .49 (28) | 1.35 (11) | (21) |
| 28 | gender | .637 (30), .155 (10) | .83 (29) | -0.06 (28) | (28) |
| 29 | MCV | .461 (26), .189 (23) | .037 (26) | -0.1 (29) | (30) |
| 30 | MCH * | .561 (29), .218 (30) | .86 (30) | -0.21 (30) | (27) |

### Composite ranking of factors based on five measures (time-to-release)

Table 2 shows the similar five rankings (and the factors are ordered by the composite ranking obtained the five) with time-to-release as the dependent variable. It is interesting that different methods do not share a common top factor: the three RSF based methods pick ferritin as the top factor, whereas Cox regression picks age, and LASSO picks calcium. When the overall rank in Table 2 is combined with the overall rank in Table 1, we have a composite rank using 10 ranking lists (last column in Table 2).

**Table 2:** Similar to Table 1 for time-to-release analyses.

| combined rank<br>(time-to-release) | var | single-var RSF<br>D(rank), IBS(rank) | Cox<br>pv(rank) | C full RSF<br>VIM$\times 10^3$ (rank) | LASSO<br>(rank) | composite10<br>(rank) |
| --- | --- | --- | --- | --- | --- | --- |
| 1 | ferritin | .327(1), .099 (1) | 1.9E-17 (7) | 21.4 (1) | (2.5T) | (8) |
| 2 | RDW.SD | .355 (4), .103 (5) | 2.6E-19 (3) | 7.87 (3) | (4.5T) | (6) |
| 3 | WBC | .361 (6), .1 (2) | 8.8E-18 (5) | 5.8 (6) | (2.5T) | (2) |
| 4 | age | .338 (2), .108(12) | 1.5E-27 (1) | 15.8 (2) | (4.5T) | (7) |
| 5 | calcium | .354 (3), .107 (9) | 3.8E-21 (2) | 4.08 (7) | (1) | (1) |
| 6 | d-dimer | .368 (7), .104 (6) | 3.6E-15 (9) | 6.56 (4) | (6.5T) | (5) |
| 7 | NEU * | .359 (5), .102 (3) | 1.1E-17 (6) | 6.48 (5) | (17.5T) | (3) |
| 8 | HGB | .39 (11), .105 (7) | 6.4E-19 (4) | 1.89 (10) | (6.5T) | (14) |
| 9 | HCT * | .395 (12), .105 (8) | 6.5E-19 (8) | 1.04 (18) | (9T) | (16) |
| 10 | LDH | .38 (9), .11 (14) | 8.9E-15 (10) | 2.85 (9) | (19) | (11) |
| 11 | urea | .373 (8), .109 (13) | 3.7E-14 (11) | 3.19 (8) | (26) | (4) |
| 12 | glucose | .397 (13), .112 (17) | 8.7E-8 (16) | 1.52 (11) | (11) | (9) |
| 13 | BAS | .42 (17), .103 (4) | 0.64 (29) | 1.35 (15) | (15.5T) | (13) |
| 14 | MCHC | .441 (21), .118 (26) | 8.1E-9 (14) | 1.4 (13) | (9T) | (21) |
| 15 | LYM | .41 (15), .107 (11) | 1.4E-4 (20) | 0.81 (19) | (21) | (12) |
| 16 | RDW.CV * | .422 (18), .114 (20) | 6.7E-10 (12) | 1.42 (14) | (27) | (18) |
| 17 | gender | .541 (30), .107 (10) | 0.002 (23) | 0.78 (20) | (9T) | (25) |
| 18 | creatinine * | .387 (10), .115 (23) | 4.3E-9 (13) | 0.43 (21) | (24) | (10) |
| 19 | sodium | .415 (16), .117 (25) | 1.5E-5 (18) | 0.32 (23) | (12.5T) | (15) |
| 20 | potassium | .409 (14), .115 (22) | 7.7E-5 (19) | 0.29 (25) | (17.5T) | (17) |
| 21 | MCV | .454 (26), .118 (27) | 3.7E-6 (17) | 1.05 (17) | (15.5T) | (29) |
| 22 | PLT | .452 (25), .112 (16) | 0.217 (27) | 1.1 (16) | (20) | (27) |
| 23 | MPV | .467 (27), .119 (28) | 8E-8 (15) | 0.37 (22) | (14) | (24) |
| 24 | EOS | .48 (28), .115 (24) | 0.777 (30) | 1.48 (12) | (12.5T) | (19) |
| 25 | PDW | .44 (20), .114 (19) | 4.7E-4 (21) | 0.13 (28) | (22.5T) | (26) |
| 26 | MON | .447 (24), .111 (15) | 0.09 (26) | 0.24 (26) | (22.5T) | (28) |
| 27 | ALT | .445 (23), .112 (18) | 0.002 (24) | 0.21 (27) | (25) | (23) |
| 28 | AST * | .425 (19), .114 (21) | 0.0014 (22) | 0.12 (30) | (30) | (20) |
| 29 | chloride | .444 (22), .12 (29) | 0.03 (25) | 0.1 (29) | (29) | (22) |
| 30 | MCH * | .509 (29), .122 (30) | 0.22 (28) | 0.31 (24) | (28) | (30) |

To compare the time-to-death and time-to-release obtained ranking, we plot the two 1/ranks versus the composite-19 rank in Fig.1(A). Generally speaking, the two are consistent. When the two are less consistent, a “bubble” is formed. We mark the name in black if a factor is ranked higher (by more than 3) in the time-to-death analyses, and in blue if the factor is ranked higher in the time-to-release analysis (and grey if the ranks are similar). We can see that neutrophils, urea, glucose, creatine, lymphocytes, etc. are ranked higher in time-to-death runs, whereas RDW.SD, age, ferritin, HGB, etc. are ranked higher in time-to-release runs.

**Figure 1:**
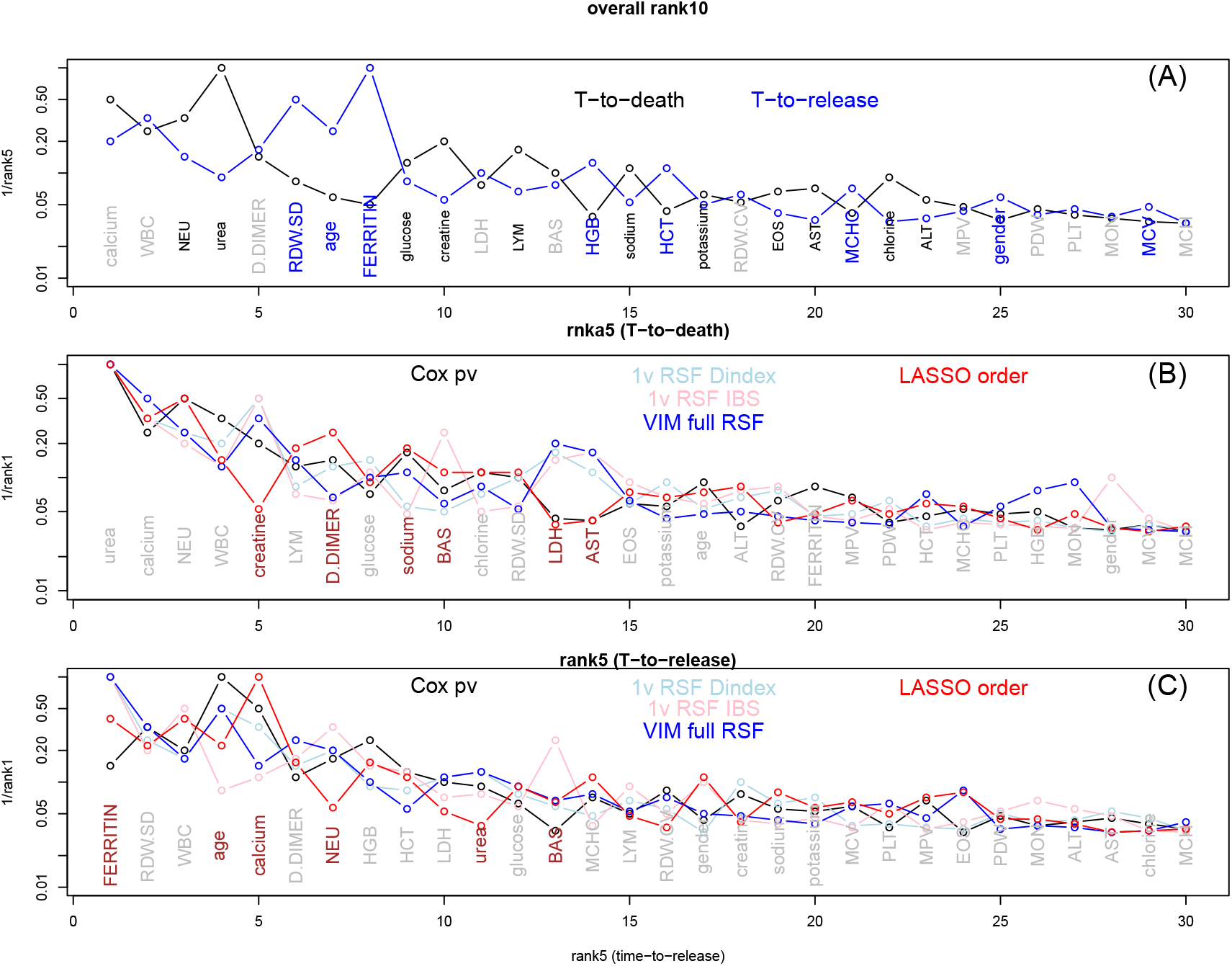
(A) Comparing the composite rank based on 5 time-to-death analyses (black) and the composite rank based on 5 time-to-release analysis (blue). The x-axis is the composite rank based on 10 analyses, and y-axis is 1/(composite rank using 5 analyses). (B) Comparing the five ranks obtained from five time-to-death analysis. The x-axis is the composite rank and y-axis is 1/(individual rank). (C) Similar to (B) for ranks from five time-to-release analyses.

Figure 1(B) and (C) also show if the individual ranks within the time-to-death group and those within the time-to-release group are consistent or not. If a factor has a large variance (normalized by the mean rank) among individual ranks, it is marked by the brown color, otherwise by grey color. All curves in Fig.1 are decreasing functions, indicating a general agreement among all ranking lists.

### Correlated factors

Because collinearity in a regression model is a problem of concern, we examine variable pairs that are correlated with each other. We use plotting of the raw data, correlation coefficient, p-value for testing correlation to be zero, the *R*^2^ from regression to determination the correlation status. Several issues are considered: (1) we check the deceased samples and survived samples separately; (2) we check both the original data and log-transformed data; and for the same reason, the correlation coefficient and testing zero correlation is based on the non-parametric Spearman method; (3) if the visual impression of the scatter plot is a guide, the *R*^2^ from regression provides a better quantity to use than, e.g. p-value for testing zero correlation.

We found six pairs of strongly correlated variables: urea and creatine, neutrophils and white blood cell, AST and ALT, RDW.CV and RDW.SD, HGB and HCT, MCV and MCH. The measure of their correlation is shown in Table 3. The lower ranked factor of a pair is marked with asterisk in Tables 1 and 2. There are more correlated variable pairs than those shown in Table 3, e.g., sodium and chloride. We use a more conservative *R*^2^ cutoff point, and require the correlation in both survived and deceased group.

**Table 3:** Factor pairs that have very strong correlation (with *R*^2^ > 0.6 in both survived and deceased group, either in the original level or log-transformed level). The correlation coefficient (cc) and p-value for testing cc=0 both refer to Spearman correlation.

| factor1 | factor 2 | deceased samples: n, $R^2$(linear)/(log),<br>cc(Spearman), pv(Spearman) | survived samples: n, $R^2$(linear)/(log),<br>cc(Spearman), pv(Spearman) |
| --- | --- | --- | --- |
| urea | creatine | 94, 0.58/0.61, 0.78, 9.7E-21 | 308, 0.42/0.61, 0.57, 1.6E-27 |
| NEU | WBC | 89, 0.99/0.98, 0.99, 6.8E-78 | 257, 0.87/0.98, 0.91, 1.4E-97 |
| AST | ALT | 94, 0.62/0.73, 0.79, 6.5E-21 | 307, 0.6/0.73, 0.71, 4.6E-48 |
| RDW.CV | RDW.SD | 94, 0.61/0.62, 0.77, 2.3E-19 | 290, 0.66/0.62, 0.71, 9.3E-46 |
| HGB | HCT | 94, 0.96/0.96, 0.98, 4.9E-64 | 290, 0.96/0.96, 0.98, 1.1E-210 |
| MCV | MCH | 94, 0.81/0.82, 0.81, 2.1E-23 | 290, 0.86/0.82, 0.92, 1.5E-122 |

### Estimation of the number of independent factors that achieve the optimal prediction performance

Although we have the composite ranking order of factors (both composite-5 in Table 1 and Table 2 and composite-10 in the last column of Table 2), there is still a question of where to cut the list to select the relevant factors. Towards this, we use a stepwise variable selection, similar to that in regression (e.g. (Ryan, 2008)), but in the framework of RSF. We first need to clarify the meaning of adding or removing a variable. There are two versions: the first is actually add a variable starting from an empty field, or remove a variable starting from a full model. The second version is to keep all variables, but instead of an empty field with no variable, the null model refers to all variables being value-shuffled. Therefore, adding the first variable is to retain its values while keeping other variables scrambled. The difference between the two versions might be written as (up to step-*i* of a variable selection):

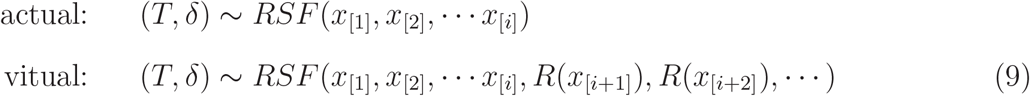

where the subscript [*i*] refers to the *i*-th variable selected, and operation R refers to random value-shuffling.

Figure 2 shows the OOB error IBS as a function of variable selection with this variable selection criterion (to stage-*i*):

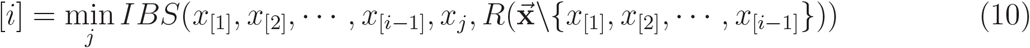

where the index *j* goes through all variables not already selected in stage-1,2, · · ·, *i* − 1, and all the rest of variables not selected remain to be value-scrambled. We have run the stepwise variable selection three times each for both time-to-death RSF and time-to-release RSF. The horizontal line is the mean of IBS from 500 runs of the full model, and dashed lines are one standard deviation from the mean (for time-to-death data, IBS= 0.0945 ± 0.000514, and for time-to-release data IBS=0.0753 ± 0.000246).

**Figure 2:**
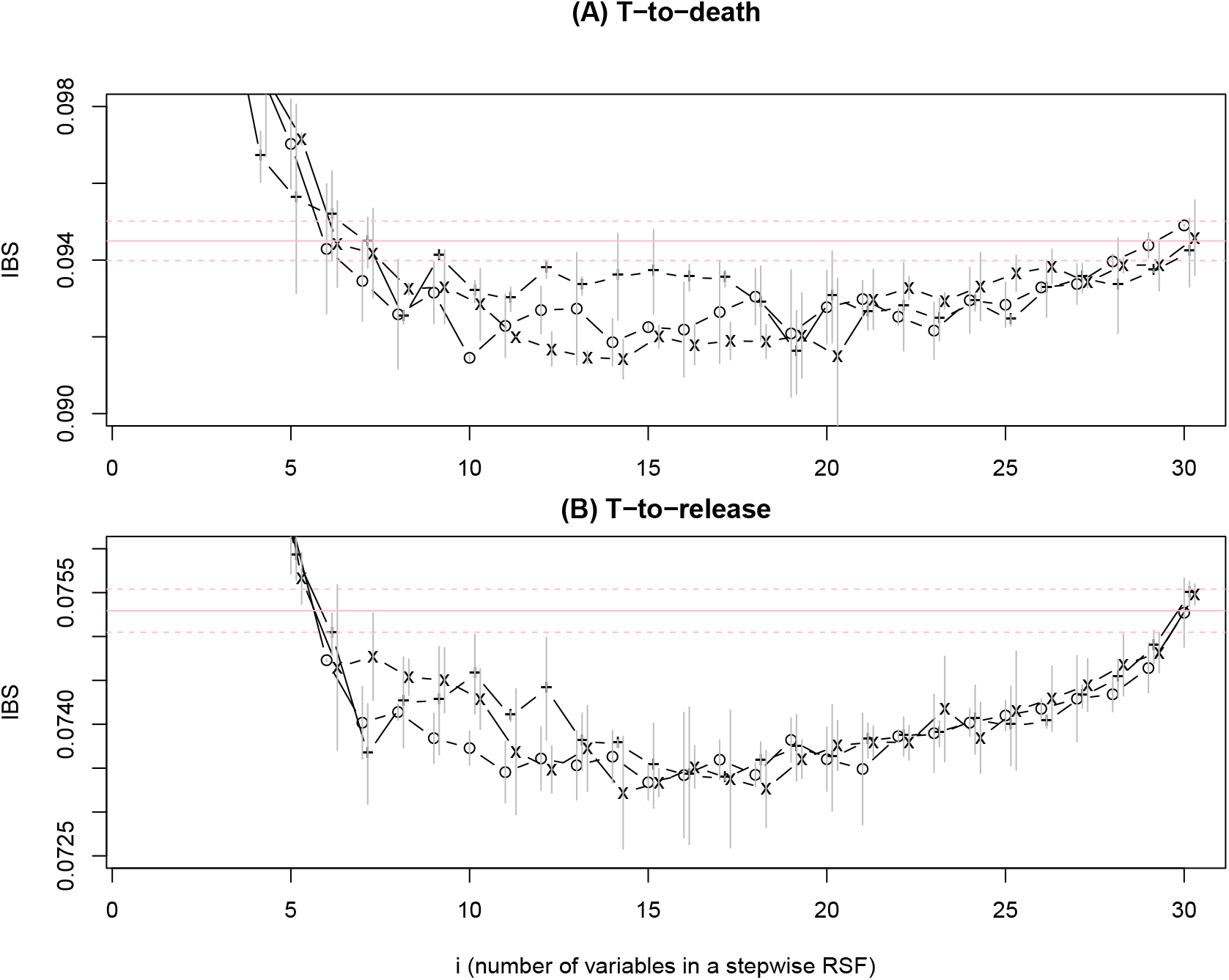
IBS from OOB samples in RSF run with the stepwise variable selection (Eq.(10)), for time-to-death data (A) and time-to-release data (B). For each variable at each stage, 10 RSF runs were carried out. The variable with the lowest mean IBS is selected, and its mean the one standard deviation up or down are shown in a vertical bar. The whole process is repeated three times (for (A) and for (B) separately). The larger IBS’s with few variables (*i* < 5) are cut off in order to zoom in the middle range of *i*’s.

There are several observations from these runs: (1) The full model is not the best performing model. It is related to a long debate on whether RF (or RSF here) needs variable selection or not (Díaz-Uriarte and De Andreés, 2006; Li, 2006). The Fig.2 shows that variable selection (reduction from the full model) is still needed for RF/RSF. However, we did not show the full range of IBS; and if we do, it will be seen that the problem of overfitting from the full model is less severe compared to other methods. (2) At each stage-*i*, multiple variables may have very similar IBS’s and the selection of *x*_[*i*]_ by Eq.(10). As a result, which particular variable is selected at stage-*i* may change from run to run (with the exception of perhaps the first few stages). Therefore, we cannot use this procedure to select risk factors. (3) On the other hand, because the three runs (per subplot) all reach the optimal performance in the middle, we can use Fig.2 to roughly estimate the number of (independent) factors to achieve the best performance. This estimation will not be precise because different runs exhibit variations; However 10∼15 (10 from Fig.2(A) and 15 from Fig.2(B)) factors should be a correct range.

### Final selection of list of risk factors

Because the factor order in Fig.2 changes from run to run, we use the pre-determined rank order (column 1 in Tables 1 and 2) to check how error decrease, i.e., the *i*-th variable added in stage-*i* is simply the rank-(*i*) variable in the ranking list: either the composite ranking order based on 5 time-to-dead analyses or on 5 time-to-release analyses. We also use both IBS and D-index as a measure of OOB prediction errors. Furthermore, both the virtual and actual variable addition were used. The resulting error curves are shown in Fig.3 (top: time-to-death runs, bottom: time-to-release runs; left: D-index, right: IBS; black: virtual addition of variables, red: actual addition of variable). It is also possible to remove the least important (low ranking) variables first (going through the ranking list backward), but the results were very similar (not shown).

**Figure 3:**
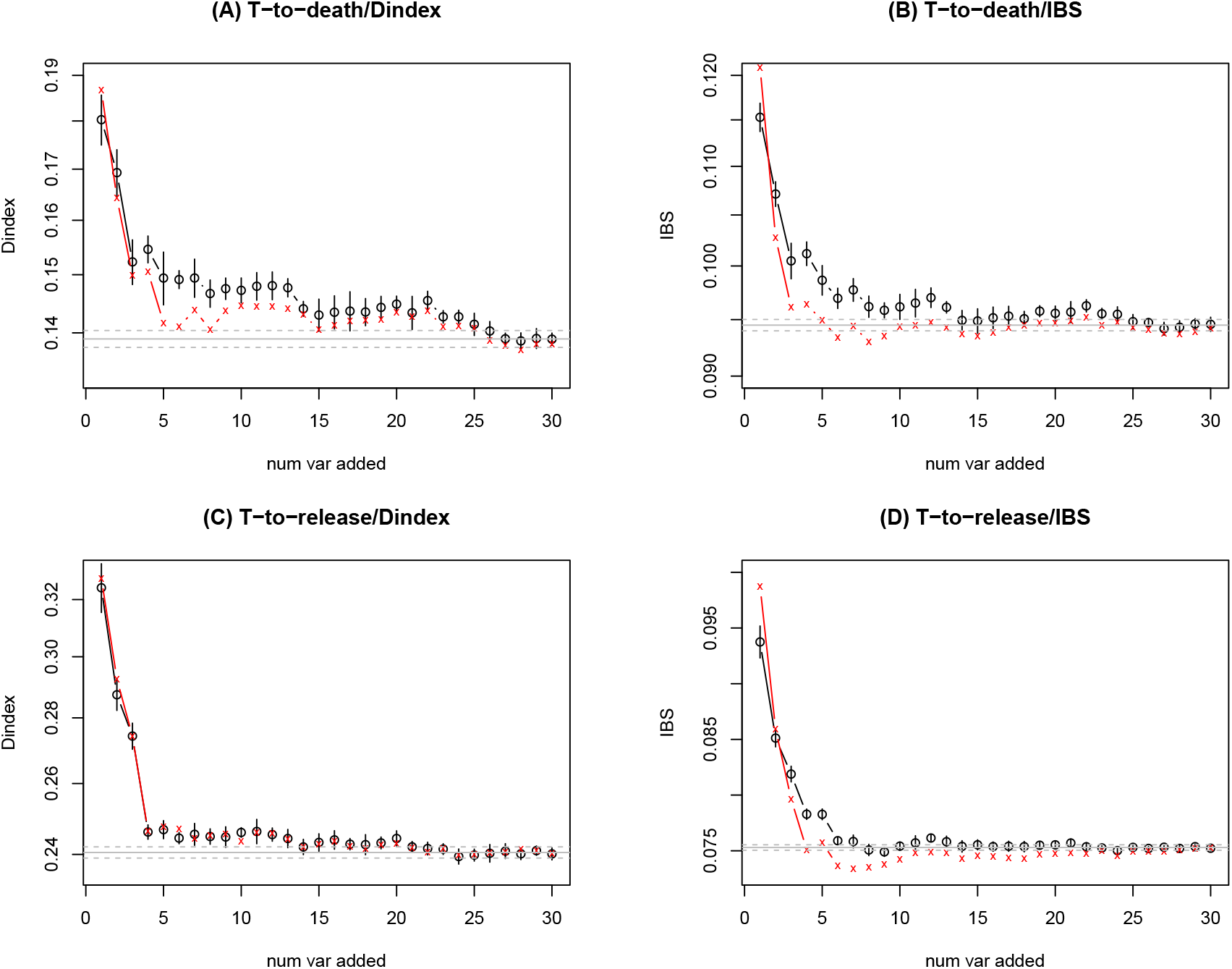
OOB RSF error (D-index on left, IBS on right) for time-to-dead (top) and time-to-release (bottom) data, as a function of *i* (stage-*i* of addition of the top-*i* ranked factors), with black for virtual variable addition and red for actual addition. The horizontal line is the mean and one standard deviation away from the mean of the full model errors (from 500 runs). The vertical bar represents one standard deviation away from the mean at stage-*i* by 10 runs.

It has been suggested in the literature that IBS is a better measure than D-index because it is more practical (Longato et al., 2020) and more quantitative (Kattan and Gerds, 2018). We also prefer the virtual variable addition over actual one because the change in the error curves in Fig.3 is smoother. Therefore, we use the black curve in Fig.3(B) to choose the top 14 factors as removing (the number 15 factor does not visibly reduce the error further, and that in Fig.3(D) to choose the top 8 factors (again because removing the number 9 factor in Fig.3(D) does not seem to reduce the error greatly). Using the similar error curve from the ranking order from VIM in full RSF model (columns 5 in Tables 1 and 2) (not shown), we will have a similar list of selected top factors. All these information and more are presented in Table 4.

**Table 4:** The risk factor selection worksheet. T2D/T2R: time-to-death/time-to-release data. VIM/rank5: rank order from variable importance of the full RSF model (column 5 of Tables 1 and 2)/composite rank order (column 1 of Tables 1 and 2). If a factor is selected by either being among the top ranking variables by the corresponding error curve, or by the p-value < 0.001 in Cox regression, it is marked by +.

| rank | factor | VIM/T2D | rank5/T2D | VIM/T2R | rank5/T2R | Cox/T2D | Cox/T2R |
| --- | --- | --- | --- | --- | --- | --- | --- |
| 1 | calcium | + | + | + | + | + | + |
| 2 | WBC | + | + | + | + | + | + |
| 3 | NEU (→WBC) | + | + | + | + | + | + |
| 4 | urea | + | + | + |  | + | + |
| 5 | d-dimer |  | + | + | + | + | + |
| 6 | RDW.SD |  | + | + | + | + | + |
| 7 | age |  |  | + | + | + | + |
| 8 | ferritin |  |  | + | + | + | + |
| 9 | glucose | + | + | + |  | + | + |
| 10 | creatine (→urea) | + | + | | | + | + |
| 11 | LDH | + | + | + |  |  | + |
| 12 | LYM | + | + |  |  | + | + |
| 13 | BAS |  | + |  |  | + |  |
| 14 | HGB | + |  | + | + |  | + |
| 15 | sodium | + | + |  |  | + | + |
| 16 | HCT (→HGB) | + | | | | | + |
| 17 | potassium |  |  |  |  | + | + |
| 18 | RDW.CV (→RDW.SD) | | | + | | + | + |
| 19 | EOS |  |  | + |  | + |  |
| 20 | AST | + | + |  |  |  |  |
| 21 | MCHC |  |  | + |  |  | + |
| 22 | chloride | + | + |  |  | + |  |
| 23 | ALT (→AST) | | | | | | |
| 24 | MPV |  |  |  |  | + | + |
| 25 | gender |  |  |  |  |  |  |
| 26 | PDW |  |  |  |  |  | + |
| 27 | PLT |  |  |  |  |  |  |
| 28 | MON | + |  |  |  |  | + |
| 29 | MCV |  |  |  |  |  |  |
| 30 | MCH (→MCV) | | | | | | |

In Table 4, we mark factors being selected by Fig.3(B), Fig.3(D), and those by two other error curves not shown. The last two columns in Table 4 mark factors which would have been selected by the standard p-value approach (p-value *<* 0.001). In fact, the threshold for p-value can be 0.01, or 0.005, and 0.001, but we consider the threshold 0.001 to be a good choice (see, for example, (Colquhoun, 2017; Ioannidis, 2018; Li et al., 2021)). If we choose factors that contribute to a better prediction performance, 21-22 factors would be selected, 17-18 of them are independent. These are the factors above the horizontal partition line, except potassium. We may consider chloride a borderline choice as it ranks last in our list, and monocyte as a borderline possibility. Note that the Cox regression p-value based selection (at p=0.001) would select MPV and PDW, which are not on our list.

The partition of factors in Table 4 into selected and not-selected ones should be considered to be ad hoc to some extent. We are only more confident of the factors being selected based on multiple lines of evidence, whereas less confident for the factors not selected. It does not imply that those not selected here are never relevant to COVID-19 severity/mortality, only that in our data they do not have the fullest level of evidence support.

## Discussions

In this work, we have carried out a careful analysis from a single dataset to determine which factors contribute to either faster death or faster release from hospital. By a literature search, we found all our selected factors were addressed in other studies and were shown to be significantly associated with the COVID-19 disease severity. Here is a partial summary:

- Low calcium level is associated with poor prognosis of COVID-19 (Di Filippo et al., 2020; Liu et al., 2020; Lee et al., 2022), probably due to its role in viral infection and replication.
- Higher neutrophil/lymphocyte ratio (NLR) is known to be associated with severity of COVID-19 (B Zhang et al., 2020; Ma et al., 2020; Ulgen et al., 2021). Individually, higher neutrophils (Reusch et al., 2021) and lymphopenia (lower lymphocyte) (Tan et al., 2020; Tavakolpour and Rakhshandehroo, 2020) both are linked to severity. Besides blood test, proteomics based neutrophil signatures have been proposed (Meizlish et al., 2021). The pathologic effect of innate immune system represented by a high level of neutrophils was investigated in (Sinha et al., 2022).
- Although leukocyte count may be normal or decreased in COVID-19 patients, the incidence of leukocytosis (higher WBC) increases in ICU patients, and has been reported to be associated with COVID-19 severity (Huang et al., 2020; Sayad et al., 2020). There is also an investigation of the causal role of WBC in disease severity by Mendelian randomization (Sun et al., 2021). Note that leukocytosis could also be related to bacterial infections, corticosteroids use, or age, and others unrelated to COVID-19 severity.
- The urea and creatine level has been observed to be higher in COVID-19 patients, and these are related to kidney abnormalities/failure, (Cheng et al., 2020; Ye et al., 2021). A recent study indicated that SARS-Cov-2 infection of kidney was possible (Jansen et al., 2022).
- D-dimer, as a byproduct of fibrinogen degradation, is closely related to thrombosis. D-dimer has been recommended as a quantity to monitor in COVID-19 patients (Thachil et al., 2020) due to ample evidence of its association with disease severity (Tang et al., 2020; Berger et al., 2020; Terra et al., 2022).
- The association of red blood cell distribution width with COVID-19 severity and mortality is reported in (Foy et al. 2020; C Wang et al., 2020; Wang et al., 2022).
- Age is perhaps the best established risk factor for severity and mortality, see, e.g. (Williamson et al., 2020).
- Elevated ferritin serum levels have been found to significantly correlate with COVID-19 severity, as discussed in these papers (Lin et al., 2020; Carubbi et al., 2021; Ahmed et al., 2021; Kaushal et al., 2022), among others.
- The short-term glucose has been shown to be a much stronger predictor for COVID-19 severity than the history of diabetes status (Singh and Singh, 2020; Coppelli et al., 2020; Ling et al., 2021; Cetin et al., 2021d; Guarisco et al., 2022). In a recent study, it was shown that higher glucose level (*>* 180) had increased the odds of death by 4-fold for non-diabetes, whereas only 1.8-fold increase for diabetes COVID-19 patients (Skwiersky et al, 2021).
- Lactate dehydrogenase (LDH), which was already known to be associated with poor outcome from viral infection, could predict COVID-19 severity (Henry et al., 2020; Han et al., 2020; Jin et al., 2022).
- Both basophils and eosinophils are found to be among the most dynamic cell populations during disease progression (Rodriguez et al., 2020), implicating them important roles played in anti-viral defense. Basophils are predicted to be a causal factor in COVID-19 severity (Sun et al., 2021), and the role of eosinophil is studied in (Lindsey et al., 2020; Xie et al., 2021; Tan et al., 2021),
- Hemoglobin and anemia has been discussed in their role of COVID-19 in (W Zhang et al., 2020; Hariyanto and Kurniawan, 2020; Benoit et al., 2021; Tao et al., 2021). Hematocrit (HCT) was mentioned as one of the laboratory tests for COVID-19 diagnosis and prognosis (Mertoglu et al., 2021). MCHC was among the lab test indicators to be significant for COVID-19 severity (Ballaz et al., 2021).
- On other essential minerals besides calcium, low abnormal sodium level (hyponatremia) was reported to be linked to COVID-19 severity (Tzoulis et al., 2021; De Carvalho et al., 2021b), negative association between potassium and severity was observed in (D Wang et al., 2020), while which patient population may have this correlation with hypokalemia was cautioned in (Szoke et al., 2021). It is interesting that the borderline item on our list, chloride, was not shown to be significantly associated with severity in (Lippi et al., 2020). While chloride’s link was investigated in other studies, its p-values were as good as other electrolytes (Wu et al., 2020; De Carvalho et al., 2021a).
- Liver biochemical biomarkers are associated with COVID-19 disease severity and clinical outcomes. Admission aspartate aminotransferase (AST) was shown to be higher in those requiring ICU stay (Gu et al. 2020; Mo et al., 2020; Ding et al., 2020; Aloisio et al., 2021). Having fatty liver disease can be a risk for COVID-19 severity (Pan et al., 2021).

Our “within-sample-meta-analysis” lead to a more robust conclusion concerning risk factors for COVID-19 severity. Using independent studies published by other groups, our cutoff in Table 4 may lead to close to zero false discovery rate (FP/(FP+TP)) or close to 100% precision (positive predictive value, TP/(FP+TP)).

We may still underestimate the number of risk factors for the following reason. Because the number of variables selected by Figs.2 and 3 represent the sufficient number of variables needed to achieve optimal performance, and adding more correlated/collineared variables would not decrease the error further. Among factors not selected in Table 4, several are related to platelet: PLT itself, and MPV and PDW related to platelet size. Because platelet is a key regulator of thrombosis and inflammation, both present in severe COVID-19 patients, it is a good candidate for risk factor (Barrett et al., 2021; Rohlfing et al., 2021; Delshad et al., 2021). However, it is suggested that platelet/large-cell-cell ratio (PLCR) (Daniels et al., 2021) is a better marker than MPV and PDW, and platelet-to-lymphocyte ratio (PLR) (Asan et al., 2021) is a better marker than platelet count. However, see counter conclusions in (Aydinyilmaz et al., 2021; Lippi et al., 2021).

Other factors not selected by our composite ranking in Table 4 include gender, monocyte, MCH/MCV (mean corpuscular hemoglobin/volumn). Male gender seems to be a risk for severity/mortality (Jin et al., 2022), though the effect size can be weak (Ortolan et al., 2020; Mukherjee and Pahan, 2021). We also cannot exclude the possibility of impact from gender-specific comorbidities (Ya’qoub et al., 2021). Monocytes usually make up a very small percentage of all white blood cells. Although it could play a role in COVID-19 severity (Vanderbeke et al., 2021), neutrophils with a much larger percentage in white cell population, should provide more signal. MCH/MCV tend to be less significant than other variables as reported in (Ballaz et al., 2021; Rahman et al., 2022). ALT is in a special situation because it is highly correlated with another factor selected (AST). There were indications that ALT could be a relevant biomarker by itself (Malik et al., 2021), or not (Qin et al., 2020), or within some patients (Bertolini et al., 2020).

The fact that factors not selected in our data whereas mentioned in other publications as potential relevant factors might reflect two situations. The first is that these factors are actually only weakly associated with COVID-19 severity/mortality and we are not able to detect them. The second possibility is that any two datasets can not be identical, and there are always chances that heterogeneity, unmeasured covariates, variations due to finite sample sizes may lead to inconsistent conclusions.

In the calculation of variable VIM in RSF, one variable is removed/shuffled from the full model, and the most important variable is listed first. It is tempting to extend this for a stepwise variable selection procedure by removing/shuffling the important variable at each stage (currently we remove/shuffle the least important variable first). However, this procedure does not produce an error curve that reaches plateau (result not shown). Therefore, we did not use this procedure for deciding the cutoff in our variable list.

In conclusion, with more interests in using blood test for prognosis in COVID-19 patients (COMBAT Consortium, 2022), we have carried out a careful analysis on blood-test factors affecting COVID-19 hazard in a time-to-event data from a Turkish cohort. We use multiple measures and methods to rank factors, some are traditional single-variable test (Cox proportional hazard ratio model), whereas others are multiple-variable machine learning techniques (random survival forest). A novel choice in our composite ranking is to utilize shorter hospital stay for released patients to discover protective factors, which should also be a risk factor when the factor value changes in the opposite direction. This approach complements the approach in using the time-to-death information for deceased patients. All of our top choices in the composite ranking list are confirmations to one of the other studies for risk factors for COVID-19 severity and/or mortality, resulting in a 100% positive predictive value.

## Data Availability

All data produced in the present study are available upon reasonable request to the authors

## Acknowledgements

WK thanks the support from the Robert S Boas Center for Genomics and Human Genetics.

## Notes

### Competing Interest Statement

The authors have declared no competing interest.

### Funding Statement

This study did not receive any funding

### Author Declarations

The use of data is approved by the ethics committee of the Tokat Gaziosmanpasa University.

### Summary of Updates

updated references

